# Neonatal outcomes after maternal biomarker-guided preterm birth intervention: the AVERT PRETERM trial

**DOI:** 10.1101/2023.09.13.23295503

**Authors:** Matthew K. Hoffman, Carrie Kitto, Zugui Zhang, Jing Shi, Michael G. Walker, Babak Shahbaba, Kelly Ruhstaller

## Abstract

The AVERT PRETERM trial (NCT03151330) evaluated whether screening clinically low-risk pregnancies with a validated maternal blood biomarker test for spontaneous preterm birth (sPTB) risk, followed by preventive treatments for those screening positive, would improve neonatal outcomes compared to a clinically low-risk historical population that had received usual care. Prospective arm participants with singleton non-anomalous pregnancies and no PTB history were tested for sPTB risk at 19^1/7^-20^6/7^ weeks’ gestation and followed through neonatal discharge. Screen-positive individuals (≥16% sPTB risk) were offered vaginal progesterone (200 mg) and aspirin (81 mg) daily, with twice-weekly nurse phone calls. Co-primary outcomes were neonatal morbidity and mortality, measured using a validated composite index (NMI), and neonatal hospital length of stay (NNLOS). Endpoints were assessed using survival analysis and logistic regression in a modified intent-to-treat population comprising screen-negative individuals and screen-positive individuals accepting treatment. Of 1460 eligible participants, 34.7% screened positive; of these, 56.4% accepted interventions and 43.6% declined. Compared to historical controls, prospective arm neonates whose mothers accepted treatment had lower NMI scores (odds ratio 0.81, 95% CI, 0.67-0.98, P=0.03) and an 18% reduction in severe morbidity. NNLOS was shorter (hazard ratio 0.73, 95% CI, 0.58-0.92, *P*=0.01), with a 21% mean stay decrease among neonates having the longest stays. Sensitivity analyses of the entire intent-to-treat population supported these findings. These results suggest that biomarker PTB risk stratification and preventive interventions can ameliorate PTB complications in singleton, often nulliparous, pregnancies historically deemed low risk.

## 1. Introduction

Preterm birth (PTB) remains the leading cause of perinatal mortality [1,2]. Children born prematurely are at great risk for chronic medical conditions [3,4] and developmental delays. These risks are inversely proportional to the neonate’s gestational age at birth (GAB). Survival gains over the last several decades are largely attributable to improved neonatal care [5] and antenatal corticosteroids [6]. Strategies targeting at-risk women, such as vaginal progesterone [7,8], low-dose aspirin (LDASA)[9], and focused care management [10] comprising increased patient outreach and education, have the potential for reducing PTB. Clinical risk factors include prior PTB [11] and shortened cervical length as measured by second-trimester transvaginal ultrasound [12,13], but their utility in identifying PTB risk is blunted by the fact that most individuals delivering prematurely have not had a prior PTB [14] nor a short cervix at the time of routine sonography (18-22 weeks’ gestation) [15,16].

Recent discoveries have identified candidate PTB biomarker risk predictors that are differentially expressed in pregnancies delivering prematurely compared to term births [17]. One such risk predictor, PreTRM, has been validated in independent and diverse study populations [18-21] and measures the ratio of maternal circulating insulin-like growth factor binding protein 4 (IGFBP4) to sex hormone binding globulin (SHBG) in the window of 18^0/7^-20^6/7^ weeks’ gestation [22]. Proposed biological links between these proteins and sPTB include an involvement of IGFBP4 in sensing fetal nutrient delivery and a role for SHBG in pro-inflammatory signaling within the placenta [18,19]. This predictor stratifies risk across a validated threshold [20] corresponding to twice the U.S. population sPTB risk with a sensitivity and specificity of 88% and 75%, respectively [22] and enriches for severe sPTB and PTB-associated neonatal outcomes such as hospital and neonatal intensive care unit (NICU) stays and neonatal morbidity [18-20].

There is an urgent need to identify and proactively address pregnancies at risk of PTB. The goal of the AVERT PRETERM trial (NCT03151330) was to determine whether a therapy bundle targeted to those identified at higher risk using the biomarker test, but lacking traditionally recognized risk factors, would improve neonatal outcomes. Co-primary outcomes were neonatal morbidity and mortality and neonatal hospital length of stay.

## 2. Materials and Methods

### 2.1. Study Design

The AVERT PRETERM trial compared outcomes following biomarker testing and treatment in a prospective arm with those observed in a historical control arm. The prospective arm was enrolled and followed from June 2018-September 2020 at ChristianaCare Hospital (Newark, DE), part of a regional health care system that serves a mixed urban and rural population across Delaware and Maryland. The historical control arm delivered at ChristianaCare from August 2016-July 2018.

The co-primary hypotheses posited that PTB risk stratification using the IGFBP4/SHBG test in a clinically low-risk population and focused preventive treatments for those screening positive would reduce (1) neonatal morbidity and mortality and (2) neonatal hospital length of stay compared to the historical arm.

### 2.2. Participant Recruitment

Per the inclusion criteria, the study prospectively enrolled individuals ≥18 years of age who had singleton pregnancies and no evidence of mullerian or fetal anomalies, cervical shortening (<25 mm), genetic anomalies, history of a prior PTB, cervical cerclage, or chronic maternal medical conditions with clear indications for delivery <37 weeks’ gestation. Individuals were excluded if they had a known reaction or contraindication to progesterone or aspirin. Also, due to biomarker test requirements, prospective arm participants were excluded if they had a blood transfusion during the current pregnancy, known hyperbilirubinemia, or were taking traditional or low-molecular-weight heparin.

### 2.3. Trial Procedure and Participant Management

Following consent, blood was obtained from prospective arm participants within the window of 19^1/7^-20^6/7^ weeks’ gestation, ascertained per American College of Obstetricians and Gynecologists gestational age dating criteria [23]. Samples were shipped to Sera Prognostics, Inc., (Salt Lake City, UT) and analyzed using the IGFBP4/SHBG test in a Clinical Laboratory Improvement Amendments- and College of American Pathologists-accredited laboratory, as described previously [18,24].

Test results were shared with the participant and their care provider. Participants were deemed screen positive if they had risk scores above a validated threshold [20], corresponding to ≥16.0% sPTB risk – approximately twice the sPTB risk for singleton pregnancies in the U.S. population. These individuals were offered, and consented again to receive, a preventive treatment bundle comprising daily progesterone (200 mg intravaginally), aspirin (81 mg) and care management, consisting of twice-weekly nursing contacts to monitor medication adherence and symptom development. The remainder of care was determined by the treating clinician. Individuals with risk scores <16.0% were designated as screen negative (i.e., not at higher PTB risk) and received usual obstetric care.

Baseline demographic information and medical/obstetrical history were extracted from the medical record at enrollment for the prospective arm and a validated obstetrical registry [25] for the control arm that was consistent across both the historical and prospective cohorts. Race was self-reported for all participants. Neonatal outcomes for both arms were obtained through the registry or, for prospective arm participants who delivered elsewhere, through medical record review. External data review was performed for the prospective arm and 10% of the control arm to ensure that eligibility requirements were met, with an error rate of <3% deemed acceptable in the historical arm. All PTB cases were reviewed further for accurate assessment of primary outcomes by a single investigator (MKH). As secular changes in care over the study period could affect prospective arm outcomes in a non-random fashion, major changes in guidance or management protocols were documented on a quarterly basis.

### 2.4. Outcomes

Two co-primary outcomes were selected: (1) neonatal morbidity and mortality, evaluated using a composite index (NMI); and (2) neonatal hospital length of stay (NNLOS) from time of birth to discharge. The NMI, described by Hassan et al. (2011) [7] , incorporates NICU length of stay (LOS) and is scored on a severity scale of 0 to 4, with 4 indicating neonatal death (detailed in Table S1). The Hassan multi-level NMI has been validated and used to assess neonatal morbidity in several trials [26,27]. The co-secondary outcomes were GAB and NICU LOS.

Primary and secondary hypotheses initially were tested in the prospective arm using a modified intent-to-treat (mITT) population comprising both the screen-negative participants and the screen-positive participants who consented to and initiated treatment with the interventions before 24 weeks’ gestation (Figure 1). This population was prespecified with the anticipation that not all participants would accept preventive interventions due to perceived concerns of potential risks and/or unknown benefits. The primary and secondary hypotheses were later tested in the full ITT population (all participants; Figure 1) using prespecified sensitivity analyses. Additional outcomes were evaluated using exploratory analyses. The outcomes, populations, and analysis groups used in primary, secondary and exploratory analyses are summarized in Table S2.

**Figure 1.**
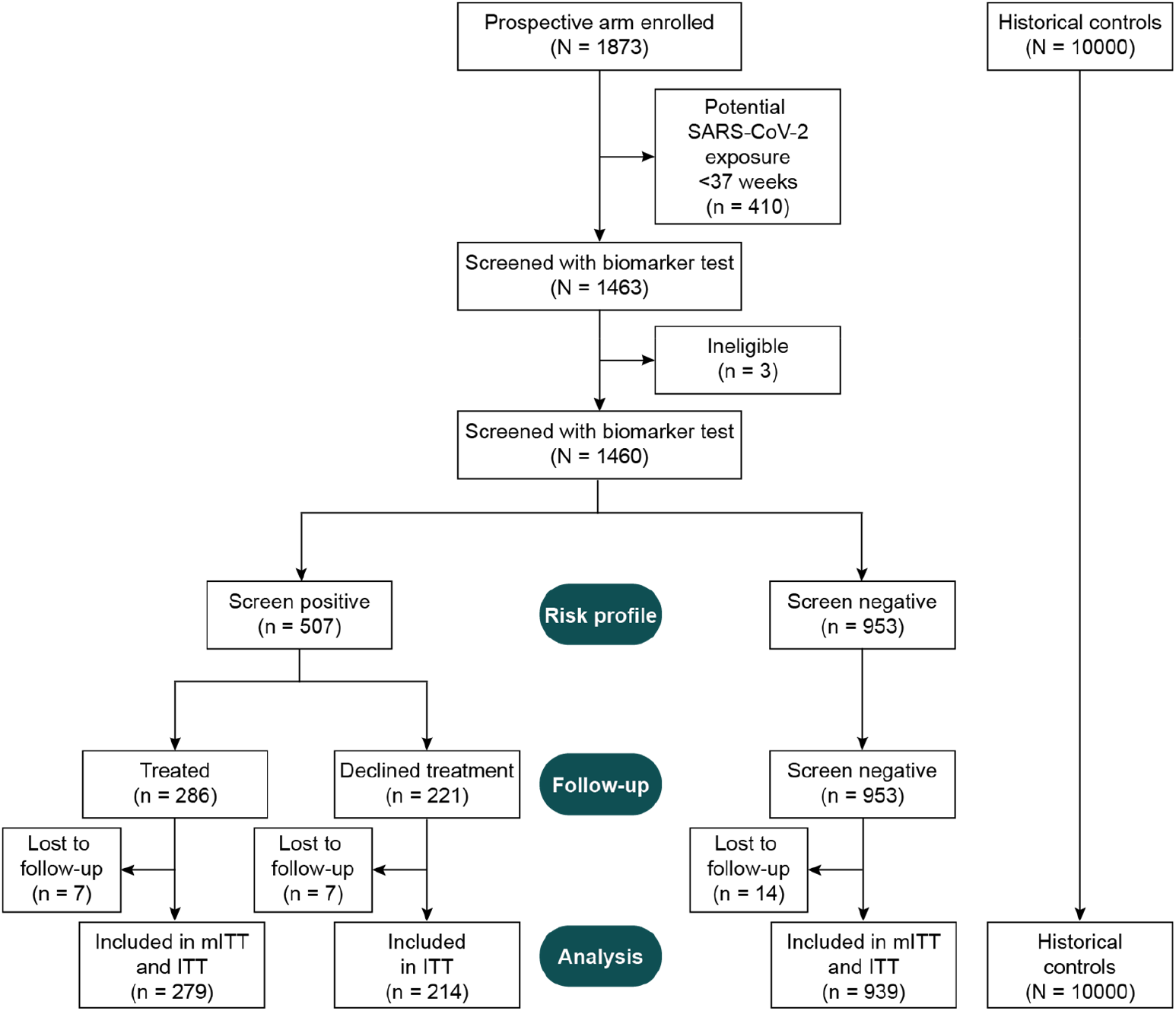
Consort diagram for study inclusion.

### 2.6. Power and sample size estimation

Pregnancy data from ChristianaCare indicated that 10000 consecutive historical controls would be available from an approximately two-year period immediately prior to study initiation, and a historical PTB rate of 9.1% was estimated. Sample size estimation was built for the co-primary outcomes using a simulated GAB distribution, the assumed singleton PTB rate of 9.1% and an effect of interventions, described elsewhere [28,29]. α-level spending of 0.05 was shared between the co-primary outcomes using Holm’s method [30].

For the NMI co-primary outcome, power was estimated conservatively using a binary comparison of the proportion of participants having NMI scores ≥3, assumed to be 2.0-2.3% in the prospective arm and near 3.6% in the control arm, based on a previous clinical utility study [31]. Assuming these proportions, with 55% compliance among screen-positive individuals, and approximately 10000 historical controls, a Fisher Exact test with a sample size of approximately 1453 individuals with outcomes in the prospective arm would provide power of 0.7-0.9 [32].

For the NNLOS co-primary outcome, the hazard ratio (HR) was expected to be 1.32-1.46, based on simulations using data from a previous clinical utility study [31]. Assuming these HRs, with 55% compliance among screen-positive participants and approximately 10000 historical controls, a Cox proportional hazards (PH) analysis with a sample size of approximately 1453 individuals with outcomes in the prospective arm would provide power of at least 0.8 [33].

### 2.7 Statistical Analysis

In April 2020, ChristianaCare Health System halted all non-COVID research. Thus, the study was stopped and the statistical analysis plan reassessed in a blinded manner. To avoid potential bias in comparing pre-pandemic historical controls to prospective participants potentially exposed to SARS-CoV-2, the plan was modified to limit the primary analysis to individuals who had reached 37 weeks’ gestation before the local spread of the virus (Figure 1).

Baseline characteristic comparisons used the Wilcoxon rank sum test to compare continuous variables between the two study arms and contingency table analysis (chi-square) to compare categorical variables, with significance set to *P*<0.05. Prespecified covariates in the primary and secondary analysis models included maternal age, parity, and opioid use. It was recognized during study design that care for newborns with neonatal opioid withdrawal syndrome (NOWS) would differ between the prospective and the historical arms due to implementation of the Eat, Sleep, Console approach [34]; therefore, analyses included maternal opiate use as a covariate, assessed as NOWS.

The NNLOS co-primary hypothesis and the NICU LOS and GAB co-secondary hypotheses were tested using Cox PH regression [33], adjusted for covariates. As described previously [35], it was predicted that clinical benefits of the test-and-treatment strategy among affected neonates could be masked by the large excess of healthy births unaffected by treatment. For this reason, the primary and secondary analyses of NNLOS, NICU LOS and GAB were conducted for prespecified extremes (quantiles) of the population corresponding to the longest hospital and NICU stays and earliest births. The predefined quantile was set at 1.2 times the observed PTB rate in the control arm. NNLOS and NICU LOS for neonates who expired were adjusted to the maximum observed stay plus one day. The severity of neonatal death was separately captured by its use as the highest value in the Hassan NMI index.

The NMI co-primary hypothesis was tested in the full mITT population (not a quantile) using ordinal logistic regression [36], adjusted for covariates. To examine the effect size at binary NMI cutoffs, differences between arms at each NMI level were calculated using a logistic regression model with a binary response variable and the covariates maternal age, parity and maternal opioid use.

Sensitivity analyses were used to assess treatment effects beyond the quantiles. NMI was additionally evaluated in the full ITT population and NNLOS, NICU LOS and GAB in the full mITT and ITT populations. In addition, primary and secondary hypotheses were tested with covariates other than maternal age, parity and opioid use. Exploratory analyses included evaluation of the PTB and sPTB rates at various GAB cutoffs, along with survival analyses for GAB <32 weeks’ gestation and NNLOS among neonates born <32 weeks’ gestation. Finally, the mean NICU days saved in the prospective versus control arm was calculated; this analysis used the actual length of stay for neonates who expired in the NICU.

Analyses were performed using R software version 4.2.2 [37]. The brant function from the brant package was used to test the proportional odds assumption, the cox.zph function from the survival package was used to test the PH assumption, and the forestplot package was used to generate forest plots. P-values <0.05 were considered significant. The co-primary outcome analyses used Holm’s multiple comparisons correction [30].

### 2.7 Trial Oversight

The study protocol was approved by the ChristianaCare institutional review board prior to participant enrollment. An independent data and safety monitoring board convened prior to study initiation, approved the protocol, and provided oversight of adverse events. The study was an investigator (MKH) study, wherein Sera Prognostics, Inc., provided the biomarker test (PreTRM®) and funding to coordinate study site recruitment, interventions, and IRB services. All study participants gave written informed consent to be included in the study. The final manuscript content was controlled by the principal investigator with editorial input by co-authors, and all authors accept responsibility for the accuracy and completeness of the data and for fidelity in the conduct of the trial.

## 3. Results

The AVERT PRETERM prospective arm was enrolled and followed from June 2018-September 2020 at ChristianaCare Hospital (Newark, DE). The historical control arm delivered at ChristianaCare from August 2016-July 2018.

At study termination, 1873 eligible individuals had been enrolled in the prospective arm, 1460 of whom were eligible, aligned with pre-COVID-19 criteria, and had been screened with the biomarker test (Figure 1). Of these, 34.7% (507/1460) screened positive by the test. Among screened individuals, 83.4% (1218/1460) had clinical outcomes and were either screen negative (77.1%, 939/1218) or screen positive accepting treatment (22.9%, 279/1218). Prospective arm outcomes were compared to those for 10000 consecutive historical controls selected from an approximately two-year period immediately preceding study initiation. The primary and secondary hypotheses were tested on the mITT population. Sensitivity analyses were evaluated in the ITT population.

Baseline participant characteristics and delivery data are shown in Table 1. The proportion of self-reported Black participants in both arms was 26.5%. The prospective arm was significantly older, more obese, and more likely to be multiparous, have hypertension and smoke than historical controls. Similarly, body mass index was higher in the prospective arm compared to historical controls – mostly due to higher weight, though the prospective arm was nominally taller than historical controls.

**Table 1.** Baseline characteristics and delivery data.

|  | Historical arm<br>(N = 10000) | Prospective arm<br>(N = 1218) | P* |
| --- | --- | --- | --- |
| Maternal Age |  |  | <0.001 |
| N | 10000 | 1218 |  |
| Mean (SD) | 29.6 (5.4) | 30.5 (5.7) |  |
| Gravida |  |  | <0.001 |
| N | 9954 | 1135 |  |
| Mean (SD) | 2.7 (1.7) | 2.41 (1.5) |  |
| Parity |  |  | <0.001 |
| N | 9953 | 1159 |  |
| Mean (SD) | 1.1 (1.2) | 0.9 (1.1) |  |
| Percent nulliparous [N, (%)] | 6544 (65.7) | 630 (54.4) | <0.001 |
| Number of miscarriages |  |  | 0.25 |
| N | 9953 | 1130 |  |
| Mean (SD) | 1.6 (1.1) | 1.6 (1.0) |  |
| Race<sup>†</sup> [N, (%)] |  |  | <0.001 |
| American Indian | 21 (0.2) | 1 (0.1) |  |
| Asian | 783 (7.8) | 76 (6.3) |  |
| Black | 2653 (26.5) | 322 (26.5) |  |
| White | 5634 (56.3) | 740 (61.0) |  |
| Other | 909 (9.1) | 74 (6.1) |  |
| Prepregnancy BMI |  |  | 0.04 |
| N | 9476 | 728 |  |
| Mean (SD) | 27.5 (8.5) | 28.2 (7.6) |  |
| BMI <19 kg/M<sup>2</sup> [N, (%)] | 403 (4.3) | 32 (4.4) | 0.85 |
| Height (inches) |  |  | <0.001 |
| N | 9838 | 1033 |  |
| Mean (SD) | 64.1 (2.7) | 64.48 (2.69) |  |
| Diabetes [N, (%)] | 127 (1.3) | 19 (1.6) | 0.42 |
| Opioid Use [N, (%)] | 242 (2.4) | 13 (1.1) | 0 |
| Hypertension [N, (%)] | 606 (6.1) | 105 (8.6) | <0.001 |
| Smoking [N, (%)] | 709 (7.8) | 100 (9.5) | 0.06 |
| Insurance type [N, (%)] |  |  | 0.67 |
| Government | 2969 (29.7) | 315 (28.6) |  |
| Other | 19 (0.2) | 1 (0.1) |  |
| Private | 7012 (70.1) | 787 (71.4) |  |
| Delivery type [N, (%)] |  |  | <0.001 |
| Dilation & evacuation | 0 (0.0) | 1 (0.1) |  |
| Primary cesarean delivery | 1577 (15.8) | 283 (20.2) |  |
| Repeat cesarean delivery | 1541 (15.4) | 177 (12.6) |  |
| Vaginal delivery | 6630 (66.3) | 923 (65.8) |  |
| Vaginal delivery after cesarean | 252 (2.5) | 18 (1.3) |  |

BMI, body mass index.
\*Continuous variables between the two study arms were compared using the Wilcoxon rank sum test, and contingency table analysis (chi-square) was used to compare categorical variables. Significance set to P<0.05.
<sup>†</sup>Race is based on self-report.

Both co-primary endpoints met statistical significance. Within the entire mITT population, NMI scores were significantly reduced in the prospective versus the historical arm (OR 0.81, 95% CI 0.67-0.98, *P*=0.03) (Table S3). Specifically, the probability of severe NMI (NMI ≥3) was reduced by ∼18%, across a range of covariate values (Table S4). NMI ≥1 (any impairment) decreased by 13-17%. For NNLOS, the control arm had an observed PTB rate of 7.1%; therefore, the 8.5% quantile (1.2 times the control PTB rate) of longest stays was used in the analysis. In this quantile, NNLOS was significantly reduced in the prospective arm versus the historical arm (HR 0.73, 95% CI, 0.58-0.92, *P*=0.01) (Table S3). The Kaplan-Meier plot for NNLOS (Figure 2), reflects a 21% reduction in mean NNLOS. Neither the proportional odds assumption for ordinal logistic regression nor the PH assumption for Cox regression were violated.

**Figure 2:**
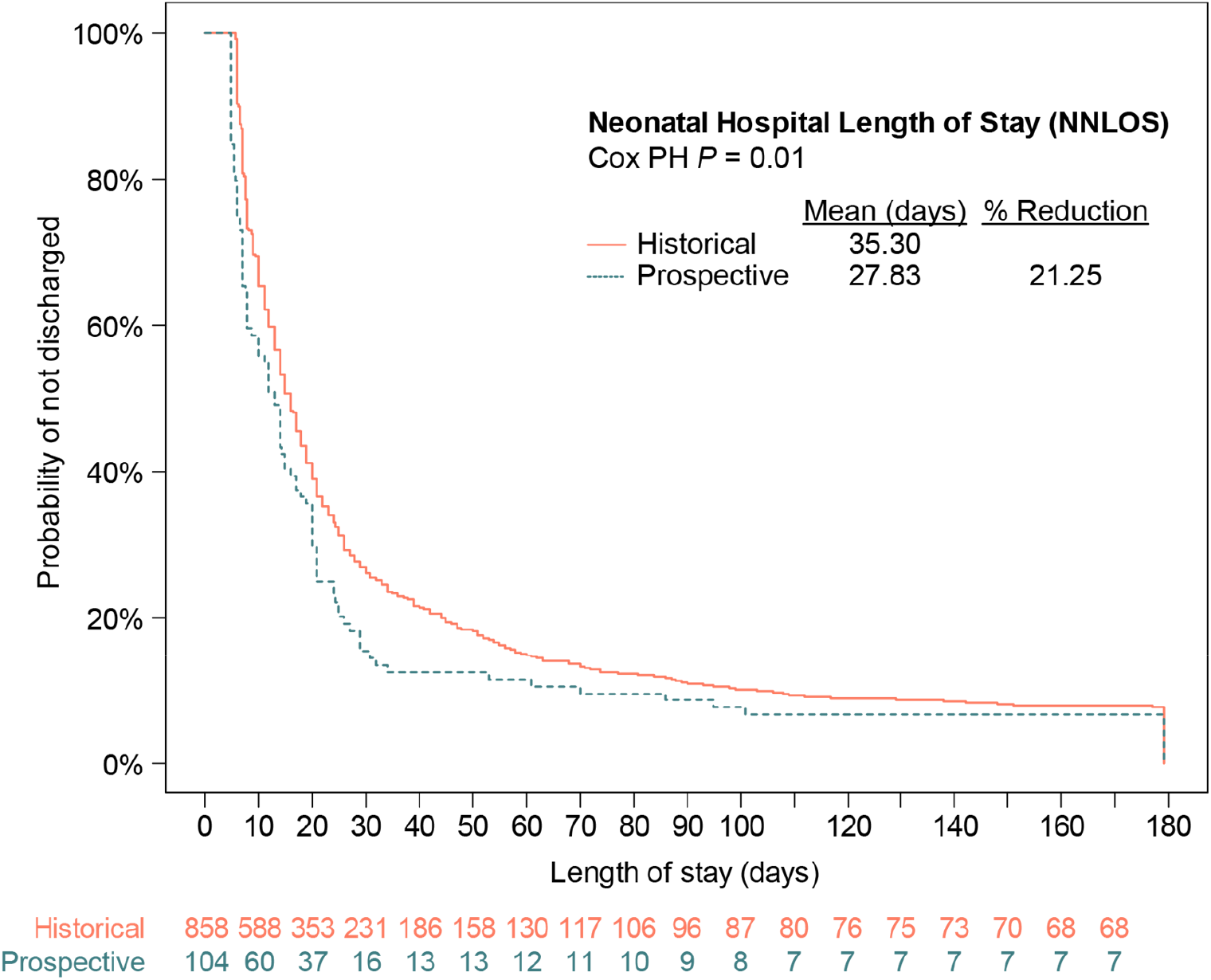
Kaplan-Meier analysis of neonatal hospital length of stay (NNLOS) for the predefined quantile of longest stays. *P* value was calculated using Cox proportional hazards (PH) regression analysis with covariates.

After statistical significance was achieved for both co-primary outcomes, the co-secondary hypotheses were tested. Neonates tended to leave the NICU earlier in the 8.5% quantile of longest stays (HR 0.83, 95% CI, 0.66-1.05), but this difference was not significant (*P*=0.12) (Table S3). No difference in GAB was observed (HR 1.04, 95% CI, 0.91-1.19, *P*=0.58) in the 8.5% quantile of earliest births (Table S3).

Sensitivity analyses of co-primary outcomes included testing with additional covariates to account for imbalances between arms. NMI and NNLOS remained significantly different between arms after adjustment for race, hypertension/preeclampsia, gravidity, and parity. In the full mITT and ITT populations, neonates were discharged from the hospital earlier than in historical controls (NNLOS; Figure 3A). NMI, significant in the mITT primary analysis, remained significantly reduced in the ITT population (Figure 3B).

**Figure 3.**
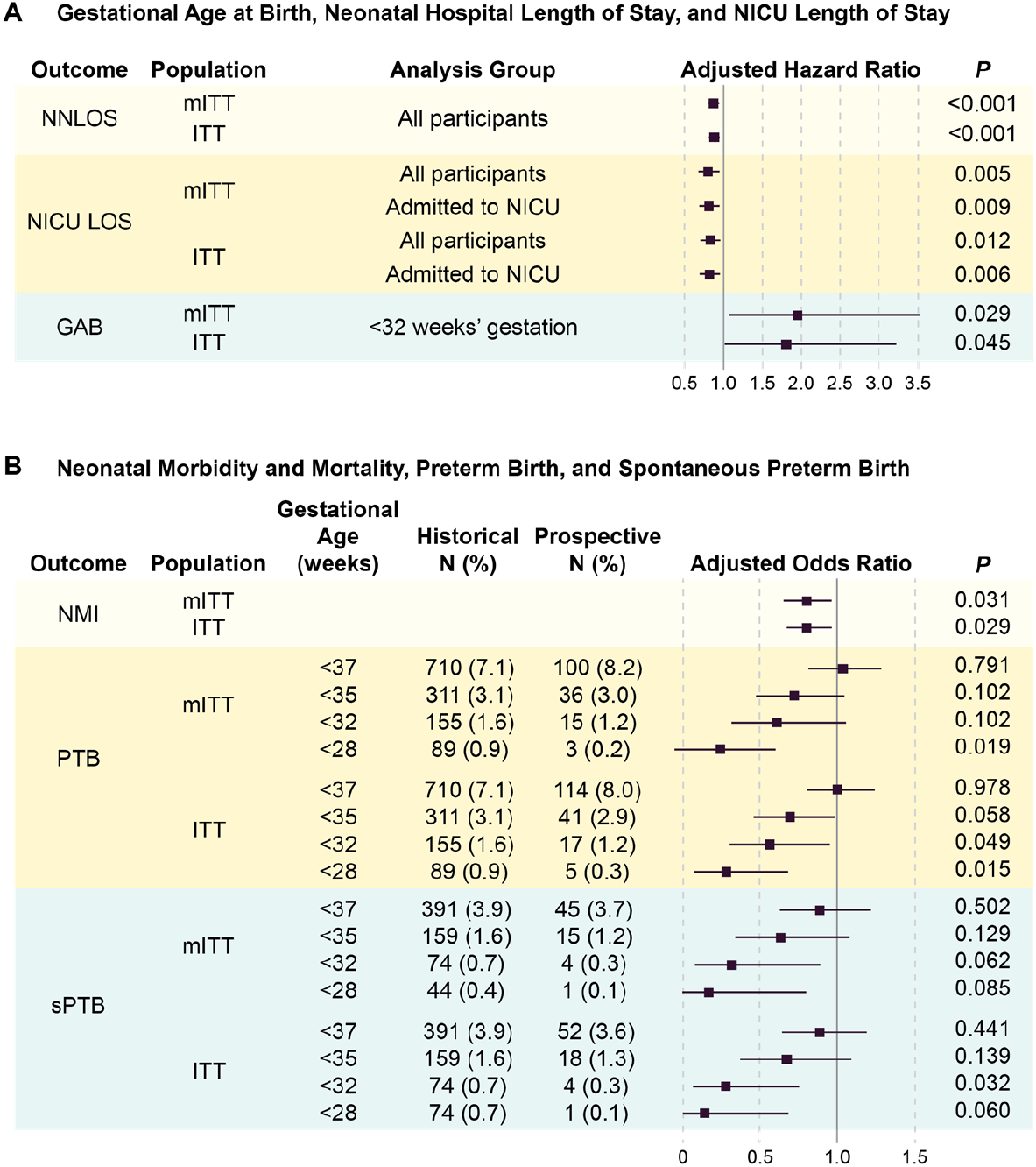
Co-primary and co-secondary outcomes in the prospective arm relative to historical controls for the mITT and ITT populations. Ratios are adjusted for parity, maternal age, and opioid use. (A) Hazard ratios below one reflect shorter neonatal hospital length of stay (NNLOS), or neonatal intensive care unit (NICU) length of stay (NICU LOS). Hazard ratios above one correspond to prolonged gestational age at delivery (GAB) relative to historical controls. (B) Odds ratios below one reflect decreased neonatal morbidity and mortality (NMI), preterm birth (PTB), and spontaneous preterm birth (sPTB) rates relative to historical controls. Solid vertical lines indicate the null value of the ratio (1.0).

Sensitivity analyses of the co-secondary outcomes showed reduced NICU LOS in the prospective arm relative to historical controls in both the mITT and ITT populations (Figure 3A), calculated to include all individuals in an arm, or only those admitted to the NICU. The mean observed NICU LOS savings across all pregnancies tested was calculated to be 0.55 days (95%CI, 0.018-1.078, *P*=0.043) in the mITT population and 0.60 days (95%CI, 0.107-1.087, *P*=0.017) in the ITT population. Mean GAB for the prospective arm was 38.5 weeks (mITT; *P*<0.001) and 38.4 weeks (ITT; *P*<0.001) versus 38.6 weeks for the historical arm.

In exploratory analyses, odds of PTB and sPTB at various gestational age cutoffs are either significantly reduced or trend in the direction of benefit in both the mITT and ITT populations (Figure 3b). Due to the decreases observed in odds of PTB, particularly for the earliest gestational age cutoffs, GAB amongst births <32 weeks’ gestation was evaluated. Although GAB was not significantly different in the secondary analyses, mean GAB for deliveries <32 weeks’ gestation is prolonged in the prospective versus the historic arm in both the mITT and ITT populations (Figure 3A). Survival analysis illustrates the GAB shift (HR, 1.94, 95%CI, 1.07-3.52, Cox PH *P*=0.029) (Figure 4A), corresponding to a 2.5-week extension of mean gestation (29.93 and 27.46 weeks for prospective and historical arms, respectively). Prospective arm neonates born <32 weeks’ gestation left the hospital earlier than did those in the historical arm (HR, 0.54, 95% CI 0.30-0.99, Cox PH *P*=0.046), with mean NNLOS differences of approximately 30% (mean of 68.47 and 97.23 days for prospective and historical arms, respectively) (Figure 4B).

**Figure 4:**
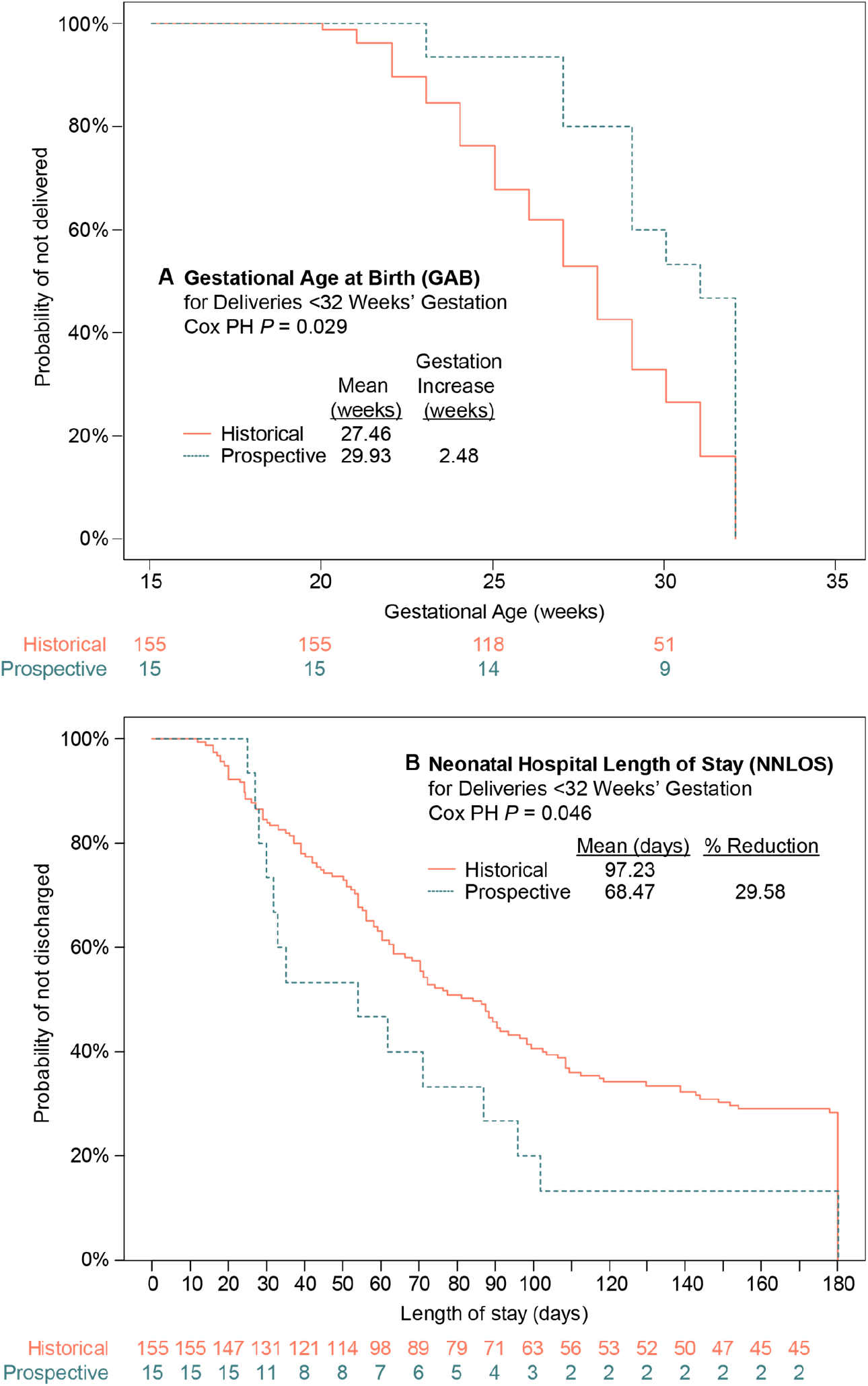
Kaplan-Meier analyses for (A) gestational age at birth (GAB) and (B) neonatal hospital length of stay (NNLOS) for neonates delivered <32 weeks’ gestation in the mITT population. *P* values were calculated using Cox proportional hazards (PH) regression analysis with covariates.

## 4. Discussion

Evidence indicates that treatment with vaginal progesterone provides benefit for individuals defined as higher PTB risk based on clinical factors [7,8]. By extension, it is reasonable to postulate that biomarker-based PTB risk stratification of otherwise low-risk pregnancies can identify individuals who might benefit from bundled interventions. The AVERT PRETERM trial results demonstrate that biomarker-based PTB risk screening and treatment with vaginal progesterone, LDASA, and care management resulted in deliveries with shorter neonatal hospital stays and less severe neonatal morbidities compared to a large historical arm, after controlling for population differences. Clinically, the impact appears greatest amongst pregnancies delivering <32 weeks’ gestation (spontaneously, or inclusive of medical indication), which remain the primary driver of newborn and child morbidity and mortality. This resonates with prior reports indicating that the IBP4/SHBG biomarker more strongly stratifies early PTBs, the most extreme health outcomes, and that stratification is not limited to PTBs arising spontaneously [18-20].

Screening a broad population of otherwise low-risk individuals for biomarker-based PTB risk and proactively treating those at higher risk presents data analysis challenges, as efficacy is expected to be limited to a minority of the population for several reasons. By definition, PTB occurs in a minority of the population, corresponding to the earliest quantile of the gestational age distribution (e.g., the ∼10^th^ percentile), and it is the earliest of these that drive the extreme lengths of stay (hospital and or NICU) seen in children born too soon. As discussed above, the biomarker test used in this study targets these outcomes [18-20]. Moreover, while PTB prevention strategies such as vaginal progesterone can shift GAB, evidence suggests that the shift is generally limited to the earliest gestational ages [7,38]. Routine statistical approaches that evaluate a central tendency (e.g., mean, median) would be dominated by the overwhelming majority of healthy term or near-term births and their associated short hospital or NICU stays, potentially diluting any impact on those pregnancies stratified by the biomarker test and expected to be improved upon by treatment. Quantile analysis, in contrast, has been demonstrated to uncover changes at the ends of the spectrum where standard analyses may obscure significant effects [39,40]. Application of this analysis approach in an obstetrical study [41] showed treatment for periodontal disease improved birthweight and gestational age at ends of the spectrum that were masked by routine statistical analyses.

Our results suggest that universal screening and treatment to prevent PTB complications amongst women lacking traditional clinical risk factors is an effective strategy. The results resonate with those from a previous randomized investigation [31] of 1191 women who either received biomarker test results and treatment or did not receive results. In that study, the NICU LOS due to sPTB was significantly shorter amongst those screened and treated versus those not receiving results (median 6.8 days versus 45.5 days; *P*=0.005). Evidence for a GAB shift was subsequently reported in a secondary analysis [35], though it was limited by low sample numbers. Moreover, a randomized controlled trial evaluating the clinical utility of the biomarker test (ClinicalTrials.gov, NCT04301518) is ongoing, and reporting of results is pending.

Given the substantially higher PTB rate among U.S. Black women (14.4%) as compared to that across the entire population (10.2%) [2], it is important to note that Black participants were represented in both AVERT PRETERM study arms with a proportion (26.5%) nearly double recent population estimates (13.6%) [2]. Results in this study population, along with those in two large and similarly diverse studies [18,19], indicate that the biomarker test will be applicable across the diverse U.S. population.

Study strengths include a multimodal intervention strategy to mitigate complications of premature delivery, as well as a biomarker test that has been validated in multiple cohorts. The data registry [25] used to obtain historical control data from the electronic medical record has been well validated, medical record review of participants delivering at other institutions was conducted, and all PTB cases were further reviewed by a single investigator. The study’s primary and secondary analyses focused on a mITT population that excluded participants not initiating treatment. This decision was made to account for subjects’ reluctance to accept treatment based on unknown benefit and potential risk during pregnancy. Nevertheless, primary outcomes remained favorably improved when examined in the ITT population.

Study limitations include the imbalances inherent in comparison of a prospective arm with a historical arm that differed in several maternal demographic and medical conditions. These differences were addressed through multivariable modeling but remain a potential source of bias. Even so, significant demographic differences in the prospective versus historical arm – older age, more hypertension and more smoking – likely biases the prospective arm toward increased PTB incidence, further underscoring the importance of these findings. Additionally, for LDASA, guideline changes have expanded the number of women eligible for treatment. A recent estimate suggests that most pregnant people should be counseled about taking LDASA in pregnancy [42]. One may argue that both progesterone and care management have limited evidence of efficacy in the situations studied here. However, the multifactorial etiologies of PTB make proving that either or both are effective in a broad range of individuals difficult, as an intervention may work on some etiologies but not others. The value of using this suite of interventions with clinically low-risk pregnancies, at least in theory and perhaps supported by results herein, is the potential for broader coverage of etiologies and better hope of success through multiple interventions. Finally, there is overlap among trial outcomes, as NICU LOS was included in the NMI index, and no multiple comparison adjustment of *P* values was made for co-secondary and exploratory outcomes.

### 4.1. Conclusions

Screening singleton, non-anomalous pregnancies lacking traditional clinical risk factors with a validated biomarker blood test for PTB risk prediction, then targeting preventive treatments for those with higher risk, shortened neonatal hospital stays and reduced neonatal morbidity and mortality. This test-and-treatment strategy can ameliorate PTB complications and associated medical, societal, and economic burdens in a large yet previously unidentifiable population: singleton, often nulliparous, pregnancies deemed clinically low risk.

## Data Availability

Data supporting the results presented here can be requested by emailing the corresponding author. Data will not be made available publicly or in any format that may violate a study participant′s right to privacy.

## Author Contributions

Conceptualization, Matthew K. Hoffman and Kelly Ruhstaller; Data curation, Zugui Zhang, Jing Shi and Michael Walker; Formal analysis, Zugui Zhang, Jing Shi, Michael Walker and Babak Shahbaba; Funding acquisition, Matthew K. Hoffman; Investigation, Matthew K. Hoffman and Carrie Kitto; Methodology, Zugui Zhang, Jing Shi, Michael Walker and Babak Shahbaba; Project administration, Matthew K. Hoffman and Carrie Kitto; Resources, Matthew K. Hoffman; Supervision, Matthew K. Hoffman and Carrie Kitto; Validation, Jing Shi and Michael Walker; Visualization, Matthew K. Hoffman, Jing Shi and Michael Walker; Writing – original draft, Matthew K. Hoffman, Jing Shi and Michael Walker; Writing – review & editing, Matthew K. Hoffman, Carrie Kitto, Zugui Zhang, Jing Shi, Michael Walker, Babak Shahbaba and Kelly Ruhstaller

## Institutional Review Board Statement

The study was conducted in accordance with the Declaration of Helsinki and approved by the Institutional Review Board (or Ethics Committee) of ChristianaCare Health Services (CCC#37149; initial approval obtained on 2/1/2018).

## Informed Consent Statement

Informed consent was obtained from all participants involved in the study.

## Acknowledgments

The authors wish to acknowledge the funding provided by Sera Prognostics, Inc., to conduct this independent investigation. Jennifer Logan, PhD, an employee of Sera Prognostics, Inc., provided medical writing and preparation support for the manuscript.

## Supplemental Materials

**Table S1.** Neonatal morbidity and mortality index (NMI) scoring and morbidity definitions.

| **Composite Neonatal Morbidity Index (NMI) Scale\*** |  |
| --- | --- |
| 0-to-4 scale integrating neonatal intensive care unit (NICU) length of stay | 0 = No events<br>1 = One event for (RDS, BPD, IVH Grade III or IV, any PVL, proven sepsis, or NEC) or 1-4 days in the NICU, and no neonatal mortality<br>2 = Two events or from 5-20 days in the NICU, and no neonatal mortality<br>3 = Three or more events or >20 days in the NICU, and no neonatal mortality<br>4 = Neonatal mortality |
| **Definitions of Neonatal Morbidity** |  |
| Category | Description |
| Respiratory distress syndrome (RDS) or Hyaline membrane disease (HMD) | Requires both diagnosis and oxygen therapy<br>Must include:<br>Oxygen therapy (fraction of inspired oxygen (FiO<sub>2</sub>) ≥0.40) until infant death or ≥24 hours or continuous positive airway pressure (CPAP) and<br>Clinical diagnosis of RDS or HMD |
| Bronchopulmonary dysplasia (BPD) | Treatment with >21% oxygen for at least 28 days, or Oxygen dependence after 36 weeks post-conceptional age |
| Intraventricular hemorrhage (IVH) | Determined by cranial ultrasound or computed tomography<br>Grade III intraventricular hemorrhage with ventricular dilatation<br>Grade IV intraventricular hemorrhage with ventricular dilatation and parenchymal extension |
| Periventricular leukomalacia (PVL) | Determined by cranial ultrasound<br>Any PVL<br>Cystic PVL |
| Sepsis | Must include:<br>Blood culture-proven sepsis, and<br>A clinically ill infant with infection defined as:<br>Bacterial sepsis of the newborn<br>Streptococcal sepsis<br>Severe sepsis |
| Necrotizing enterocolitis (NEC) | Stage I: Other – Suspect (treatment was observation)<br>Stage II: Clinical – Definite (treatment was medical)<br>Stage III: Surgical – Advanced (treatment was surgical) |
| Neonatal mortality | Neonatal death within 28 days of delivery |

\* Hassan SS, Romero R, Vidyadhari D et al. PREGNANT Trial. Vaginal progesterone reduces the rate of preterm birth in women with a sonographic short cervix: a multicenter, randomized, double-blind, placebo-controlled trial. *Ultrasound Obstet Gynecol.* 2011 Jul;38(1):18-31. doi: 10.1002/uog.9017. Epub 2011 Jun 15. PMID: 21472815; PMCID: PMC3482512.

**Table S2.** Outcomes, populations, and analysis groups used in the AVERT PRETERM trial.

| Analysis | Endpoint(s) | Population | Analysis Group |
| --- | --- | --- | --- |
| **Primary** | NNLOS | mITT | Quantile |
|  | NMI | mITT | All |
| **Secondary** | NICULOS | mITT | Quantile |
|  | GAB | mITT | Quantile |
| **Exploratory** | NNLOS | mITT, ITT | All, <32 weeks GAB |
|  | NMI | ITT | All |
|  | NICU LOS | mITT, ITT | All, Admitted to NICU |
|  | GAB | mITT, ITT | <32 weeks GAB |
|  | PTB | mITT, ITT | <37, <35, <32, <28 weeks |
|  | sPTB | mITT, ITT | <37, <35, <32, <28 weeks |

GAB, gestational age at birth; NMI, neonatal morbidity and mortality index score; NICU LOS, neonatal intensive care unit length of stay; NNLOS, neonatal hospital length of stay.

**Table S3.** Results of hypothesis tests for the co-primary and co-secondary outcomes.

| Co-primary endpoints |  |  |  |  |  |
| --- | --- | --- | --- | --- | --- |
|  | Hazard ratio | Lower confidence limit | Upper confidence limit | *p*<sup>‡</sup> | Reference arm |
| **Neonatal length of hospital stay\*<sup>§</sup>** |  |  |  |  |  |
| Historical vs prospective arm | 0.73 | 0.58 | 0.92 | 0.01 | Prospective |
| Maternal age | 1.00 | 0.99 | 1.01 | 0.75 |  |
| Nulliparous vs parous | 1.06 | 0.92 | 1.22 | 0.39 | Parous |
| Without vs with opioid use | 0.64 | 0.55 | 0.75 | <0.001 | With |
| **Neonatal morbidity and mortality index score<sup>¶</sup>** | Odds ratio | Lower confidence limit | Upper confidence limit | *p*<sup>‡</sup> |  |
| Prospective vs historical arm | 0.81 | 0.67 | 0.98 | 0.03 | Historical |
| Maternal age | 1.01 | 1.00 | 1.02 | 0.13 |  |
| Nulliparous vs parous | 1.72 | 1.53 | 1.93 | <0.001 | Parous |
| Without vs. with opioid use | 0.38 | 0.29 | 0.51 | <0.001 | With |
| Co-secondary endpoints |  |  |  |  |  |
|  | Hazard ratio | Lower confidence limit | Upper confidence limit | *p*<sup>‡</sup> |  |
| **NICU length of stay\*<sup>§</sup>** |  |  |  |  |  |
| Historical vs prospective arm | 0.83 | 0.66 | 1.05 | 0.12 | Prospective |
| Maternal age | 1.00 | 0.99 | 1.01 | 0.78 |  |
| Nulliparous vs parous | 1.04 | 0.91 | 1.19 | 0.57 | Parous |
| Without vs with opioid use | 0.42 | 0.31 | 0.59 | <0.001 | With |
| **Gestational age at birth<sup>†§</sup>** | Hazard ratio | Lower confidence limit | Upper confidence limit | *p*<sup>‡</sup> |  |
| Historical vs prospective arm | 1.04 | 0.91 | 1.19 | 0.58 | Prospective |
| Maternal age | 1.00 | 0.99 | 1.01 | 0.93 |  |
| Nulliparous vs parous | 1.18 | 1.08 | 1.29 | <0.001 | Parous |
| Without vs with opioid use | 1.10 | 0.88 | 1.37 | 0.41 | With |

GAB, gestational age at birth; NICU, neonatal intensive care unit; NMI, neonatal morbidity and mortality index score; NICU LOS, NICU length of stay; NNLOS, neonatal hospital length of stay.
\*NNLOS and NICU LOS are reported for individuals in the 8.5% quantile of longest stays in each arm.
<sup>†</sup>GAB is reported for the earliest 8.5% quantile of births in each arm.
<sup>‡</sup>P- values report the significance of the individual covariates listed, and outcome comparisons between the prospective and historical arms are adjusted for these covariates.
<sup>§</sup>Cox proportional hazards regression analysis.
<sup>¶</sup>Ordinal logistic regression analysis.

**Table S4.**
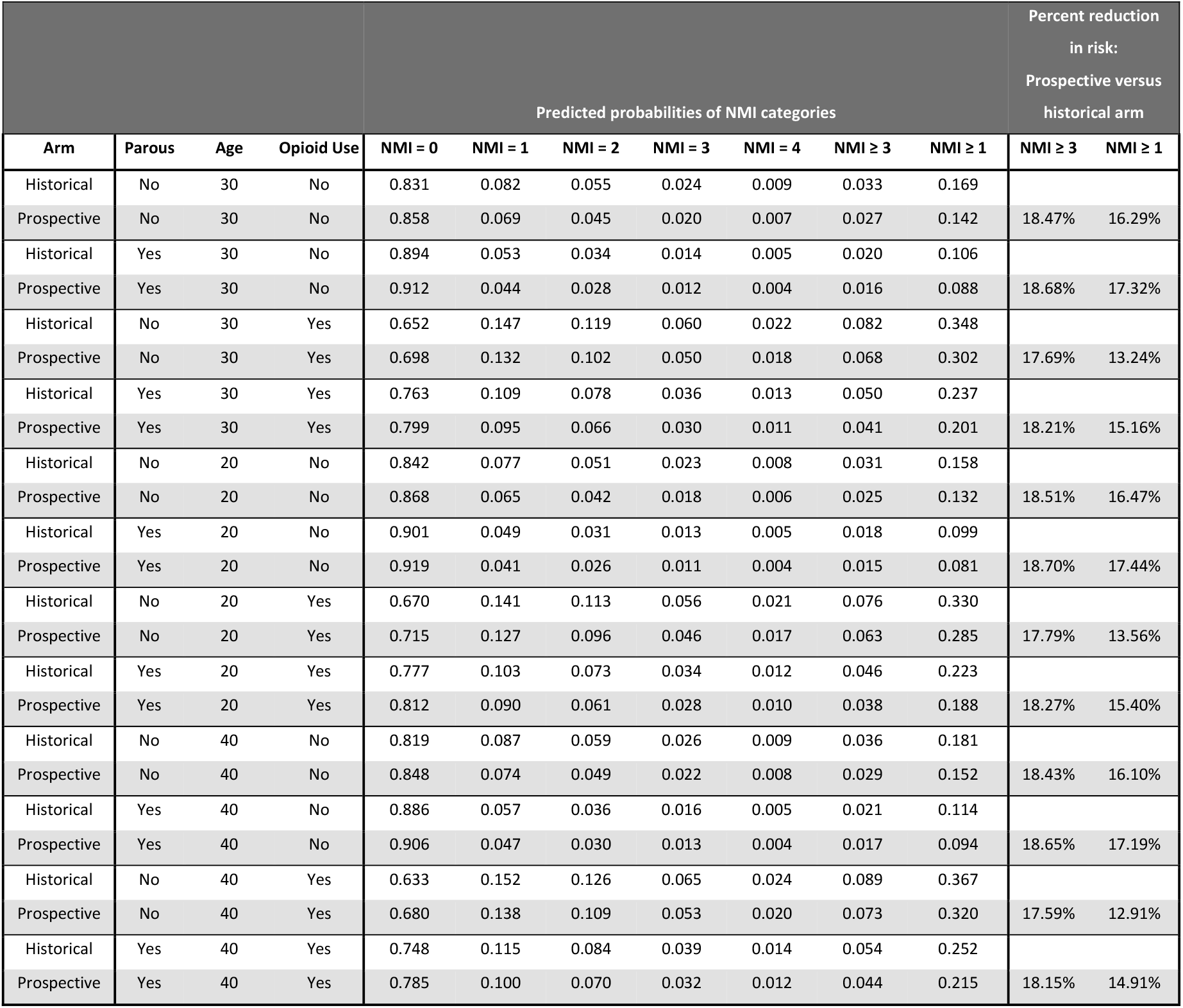
Sensitivity analysis of neonatal morbidity and mortality index score (NMI) severity level probabilities and percent differences between arms for a range of covariate values.

